# Cutaneous squamous cell carcinoma 1986 - 2019 in Germany: Incidence, Localization, Staging, and Histologic Types

**DOI:** 10.1101/2025.05.07.25327138

**Authors:** Julius Balkenhol, Thomas Dirschka, Conrad Falkenberg, Claus Garbe, Enno Swart, Lutz Schmitz

**Author notes:** Corresponding author: Lutz Schmitz. **Funding statement:** Fees for the data utilization requests at the Robert Koch Institute were funded by CentroDerm. **Data Availability:** Data used in this research is available upon a utilization request at the Robert Koch Institute. **Ethics Statement:** The study was performed under the data utilization request 5.03.04/0003#0025.

## Abstract

**Background:** Cutaneous squamous cell carcinoma (cSCC) is the second most common non-melanoma skin cancer and is associated with considerable morbidity. Population-based data analysis in Germany has largely focused on incidence and trends.

**Objectives:** To assess incidence, anatomical site- and T-stage distribution, histological subtypes of cSCC in Germany, with a focus on sex- and age-group specific patterns and regional differences.

**Methods:** A total of 213,935 first primary invasive cSCC cases diagnosed between 1986 and 2019 were analysed from four federal states of Germany with complete case ascertainment. Crude and age-standardized incidence rates (CIR, ASIR) were calculated, and subgroup analyses were performed by sex, anatomical site, histological subtype, and T-stage and region.

**Results:** CIR increased by over 500 % from 1986 to 2015, with a steeper rise in women. Incidence plateaued after 2015 in most states, except for a delayed increase in Saarland. The face (ICD-10 C44.3) was the most frequent tumor site showing equal incidence in males and females. T1 tumors predominated (88.6 %), although staging data were incomplete in 33.5 % of cases. Regional and sex-based differences were observed in both T-stage and histological subtype distribution. Spindle cell and non-keratinizing variants were associated with more advanced stages. Cancer registry data did not count more than one cSCC and carcinoma in situ such as Bowen’s disease or actinic keratosis, leading to systematic underestimation of disease burden.

**Conclusions:** cSCC incidence has risen substantially in Germany, with significant variation by sex, region, and tumor type. Improved registry protocols incorporating multiple primaries, clinical staging, and early in situ lesions are essential for accurate surveillance and healthcare planning.

**Plain Language Summary:** How common is squamous cell skin cancer in Germany and how does it behave?

We looked at cutaneous squamous cell carcinoma (cSCC), a common skin cancer that starts in the flat cells on the skin’s surface. It is the second most common skin cancer and the second most common cancer, affecting tens of thousands of people in Germany each year.

We found that the registration of new primary cSCC tumors increased more than fivefold between 1986 and 2015. However, incidence rates plateaued from 2015 to 2019. Tumors most often appeared on the face, but the distribution by site differed between men and women. Men developed cSCC at younger ages and more frequently than women. Approximately 90% of tumors were diagnosed at an early stage, but staging information was missing for about 34% of cases.

cSCC develops on chronically sun-damaged skin, and patients often have more than one tumor. There are also early skin changes that require treatment. Because cancer registries count only one tumor per person and ignore these early lesions, the true burden of treating patients with chronic sun damage is underestimated.

We concluded that cSCC has become more common in Germany, with clear differences by sex, age, and region. Improving cancer registries to record all tumors will provide a more accurate picture of stage and subtype distribution. Recognizing high-risk groups, will help guide prevention, screening, and earlier treatment strategies.

**What is already known about this topic?:** Cutaneous squamous cell carcinoma (cSCC) is the second most frequent malignancy overall as well as the second most frequent skin tumor. Epidemiological research has long focused on Basal Cell Carcinoma (BCC) and cSCC conclusively. Recent research has addressed major differences in their epidemiology, highlighting differnces in incidence rates across genders and age groups. The largest data set on squamous cell carcinoma analysed so far was 145.000 cases.

**What does this study add?:** This study provides conclusive results on incidence, localization, tumor stages and histologic types comparing men, women and age groups based on large scale data with over 200,000 primary tumors over a period of more than 30 years.

**What is the translational message?:** Identification of gender- and age-specific risk patterns in cSCC enables the formulation of targeted prevention strategies, screening recommendations, and earlier diagnosis and optimized management of high-risk populations. The burden of morbidity and tumors associated with chronic actinic damage remains underestimated in current literature and cancer statistics. Demographic changes are expected to increase the disease burden substantially, although the exact magnitude remains uncertain.

## Introduction

Cutaneous squamous cell carcinoma (cSCC) is the second most common type of nonmelanoma skin cancer (NMSC), following basal cell carcinoma (BCC), and is increasingly recognized for its clinical significance due to its invasive potential and association with morbidity, particularly in older populations ^1,2^. While BCC is three times more prevalent overall, cSCC carries a substantially higher risk of metastasis and local tissue destruction ^3,4^. Despite this, population-based epidemiological data on cSCC remain limited in many countries, including Germany ^1,5^. Epidemiological surveillance of cSCC faces several challenges. Cancer registries in Germany typically record only the first tumor per patient, excluding recurrent or multiple primaries that are common in cSCC ^1^. Moreover, precursor lesions such as actinic keratosis (AK) and Bowen’s disease, which represent early stages of UV-induced carcinogenesis, are not documented in cancer statistics. This leads to underestimation of both the true incidence and the therapeutic burden of chronic keratinocytic disease ^6^. Several European studies have demonstrated increasing cSCC incidence, with variations by sex, anatomical site, and age ^2,7,8^. However, many lack detailed stratification by histological subtype, tumor stage, and localisation. This study aims to address this gap by providing a comprehensive assessment of cSCC based on high-quality registry data from four German federal states with verified complete case ascertainment (CCA) ^9^. We evaluated trends in incidence, anatomical and histological characteristics, and staging, with particular attention to sex- and age-specific patterns, and critically discuss the implications for surveillance and healthcare planning.

## Materials and Methods

This retrospective population-based study used anonymized data from the German Centre for Cancer Registry Data (ZfKD) at the Robert Koch Institute (RKI) [9]. Data were obtained from four federal states of Germany – Schleswig-Holstein, Lower Saxony, North Rhine-Westphalia, and Saarland – with documented CCA ≥ 90% over at least five consecutive years ^1^. The observation period ranged from 1986 to 2019, depending on data availability in each region. Eligible cases included first primary invasive cutaneous squamous cell carcinomas (cSCC), defined by ICD-10 code C44 and histological subtypes coded as 8070–8084 in the ICD-O-3 system. Cases of carcinoma in situ, intraepidermal neoplasia, and multiple tumors from the same individual were excluded, as cancer registries document only the first malignancy per person and anatomical site. The stepwise case selection process is illustrated in Figure 1. Variables included sex, age at diagnosis, year of diagnosis, federal state, tumor site, laterality, histological subtype, and T-stage. Tumor sites were classified using ICD-10 topography codes C44.0 to C44.9, corresponding to specific anatomical regions: lip (C44.0), eyelid (C44.1), ear and external auditory canal (C44.2), face (C44.3), scalp and neck (C44.4), trunk (C44.5), upper limbs (C44.6), lower limbs (C44.7), overlapping sites (C44.8), and unspecified skin (C44.9). Tumors with overlapping or unspecified localisation (C44.8, C44.9) were excluded from anatomical analyses. Histological subtypes were recorded according to ICD-O-3 morphology codes. The category “not otherwise specified” (NOS) refers to cases lacking further subclassification in the pathology report and was used when no additional histological detail beyond cSCC was available. T-stage information was available for 66.5% of cases. As both clinical and histopathological parameters are required for TNM staging, missing values likely reflect incomplete information at the reporting site. Crude incidence rates (CIR) and age-standardised incidence rates (ASIR) were calculated per 100,000 person-years using the direct method, based on the 2018 European Standard Population. Population denominator data were obtained from the GENESIS database of the German Federal Statistical Office. Age-specific rates were calculated in ten-year intervals. Average annual percentage change (AAPC) was computed to evaluate temporal incidence trends. AAPC reflects the mean yearly rate of change over the full observation period and was derived using log-linear regression models. A positive AAPC indicates an increasing trend, and statistical significance was determined using 95% confidence intervals. Descriptive statistics were used to summarize tumor and patient characteristics. Continuous variables were analyzed using means and standard deviations and compared between groups using independent t-tests or one-way ANOVA. Categorical variables were reported as absolute and relative frequencies, with group comparisons performed using chi-square tests. Exact binomial tests were used to assess asymmetry in laterality distributions. A p-value *<* 0.05 was considered statistically significant. All analyses were performed using SPSS software (version 25.0, IBM Corp.). This study was approved by the Robert Koch Institute (utilization request 5.03.04/0003 #2510). All data were fully anonymized prior to analysis. Cancer reporting is mandatory in Germany, and patients are informed of their right to opt out.

**Figure 1.**
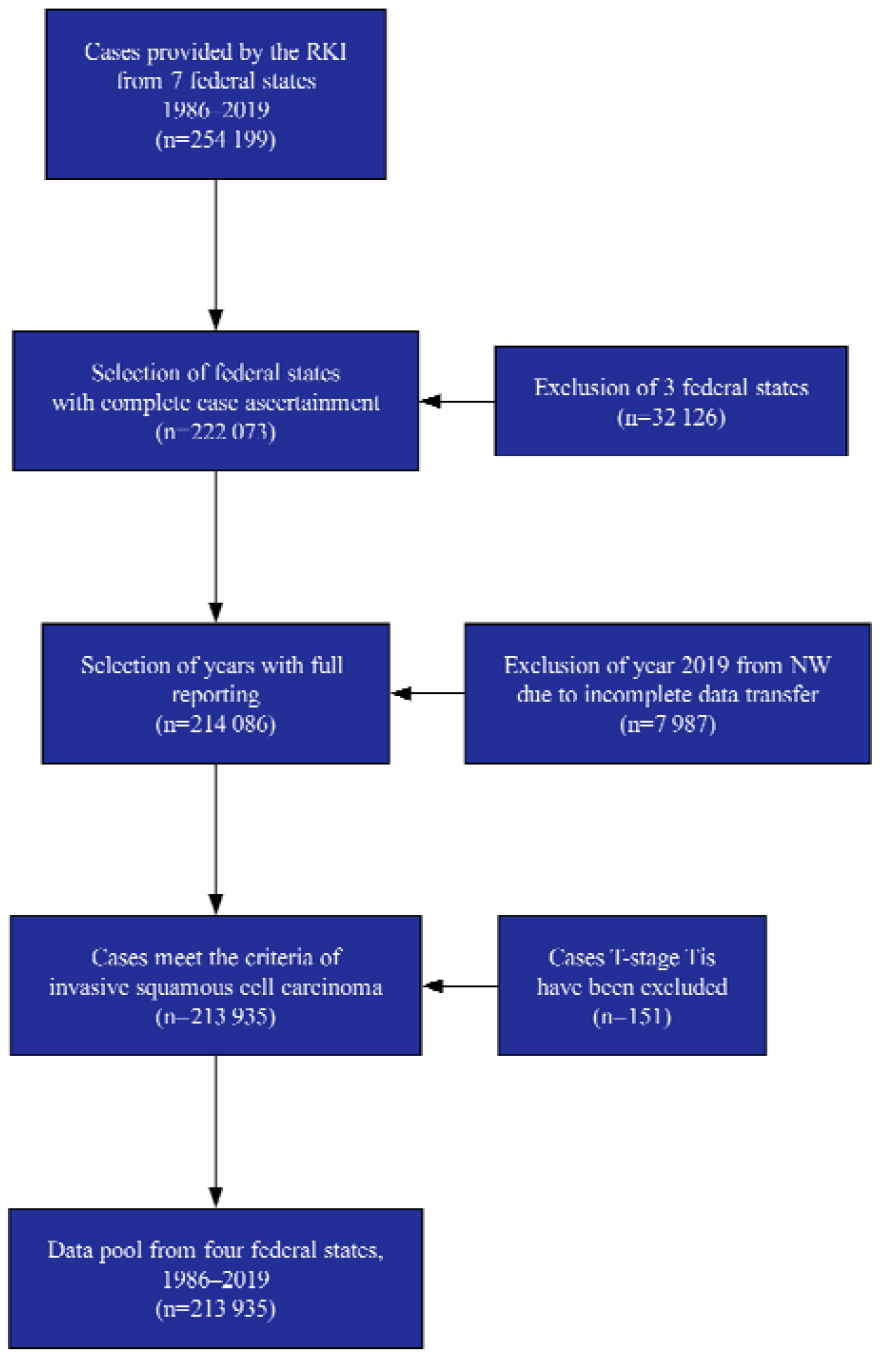
Applying exclusion criteria on the total data frame provided by the Robert Koch Institute (RKI). North Rhine-Westphalia (NW).

## Results

A total of 213,935 cases of first primary invasive cSCC were included. Of these, 59.5% occurred in men. The mean age at diagnosis was 76.9 years (SD ±10.4) (Table 1), with men diagnosed slightly earlier than women (76.2 vs. 77.9 years; p *<* 0.0001). Most cases were reported from North Rhine-Westphalia (51.4%), followed by Lower Saxony (30.6%), Schleswig-Holstein (14.2%), and Saarland (3.8%).

**Table 1.** Crude and Age-Standardized Incidence Rates by Age Group and Sex.

| Age Group | Population at Risk |  | Frequency |  | CIR |  | ASIR |  |
| --- | --- | --- | --- | --- | --- | --- | --- | --- |
|  | Females | Males | Females | Males | Females | Males | Females | Males |
| 0-9 | 19,342,66 | 20,406,805 | 8 | 5 | 0.04 | 0.02 | 0.00 | 0.00 |
| 10-19 | 22,638,89 | 23,982,419 | 14 | 14 | 0.06 | 0.06 | 0.00 | 0.00 |
| 20-29 | 25,318,07 | 26,615,778 | 72 | 87 | 0.28 | 0.33 | 0.03 | 0.02 |
| 30-39 | 27,288,17 | 27,645,001 | 305 | 311 | 1.12 | 1.12 | 0.14 | 0.11 |
| 40-49 | 33,495,95 | 33,961,452 | 1,655 | 1,808 | 4.94 | 5.32 | 0.56 | 0.44 |
| 50-59 | 32,505,30 | 32,377,226 | 4,541 | 5,968 | 13.97 | 18.43 | 2.05 | 1.29 |
| 60-69 | 26,629,35 | 24,977,780 | 10,903 | 19,201 | 40.94 | 76.87 | 6.75 | 3.22 |
| 70-79 | 23,286,00 | 18,810,777 | 27,635 | 52,654 | 118.68 | 279.91 | 17.86 | 7.41 |
| 80-89 | 13,264,14 | 7,247,220 | 31,618 | 41,013 | 238.37 | 565.91 | 15.89 | 7.08 |
| 90+ | 2,653,84 | 827,518 | 9,996 | 6,127 | 376.66 | 740.41 | 11.91 | 6.10 |

*Legend:* Age-stratified crude incidence rates (CIR) and age-standardized incidence rates (ASIR) per 100,000 person-years, based on the 2018 European Standard Population. Case frequencies and population denominators are shown separately for females and males across age groups.

Between 1986 and 2019, crude incidence rates (CIR) increased by 528.4% (Table 2). This increase was greater in women (653.0%) than in men (454.2%), with an AAPC of 6.31% versus 5.72% (p *<* 0.0001), (Figure 2). Age-standardized rates showed a similar pattern. After peaking in 2015, incidence stabilized or declined slightly in three of the four states, with Saarland exhibiting a delayed rise (Figure 3).

**Table 2.** Crude and Age-Standardized Incidence Rates Over Time with 95% Confidence Intervals.

| Year | CIR Total | 95% CI CIR | ASIR Total | 95% CI ASIR |
| --- | --- | --- | --- | --- |
| 1986 | 10.46 | 8.50–12.42 | 14.02 | 11.28–17.60 |
| 1987 | 9.68 | 7.80–11.55 | 13.98 | 11.12–17.68 |
| 1988 | 12.52 | 10.39–14.66 | 16.85 | 13.90–20.54 |
| 1989 | 14.37 | 12.09–16.64 | 18.05 | 15.15–21.64 |
| 1990 | 13.14 | 10.97–15.31 | 17.39 | 14.37–21.09 |
| 1991 | 14.67 | 12.38–16.96 | 18.33 | 15.47–21.80 |
| 1992 | 16.05 | 13.67–18.44 | 20.61 | 17.50–24.30 |
| 1993 | 15.86 | 13.49–18.23 | 20.38 | 17.31–24.00 |
| 1994 | 15.40 | 13.07–17.74 | 19.43 | 16.47–22.90 |
| 1995 | 13.37 | 11.20–15.55 | 16.76 | 14.05–19.97 |
| 1996 | 16.60 | 14.18–19.03 | 20.40 | 17.43–23.85 |
| 1997 | 15.64 | 13.28–17.99 | 17.98 | 15.32–21.07 |
| 1998 | 16.57 | 14.14–19.00 | 19.52 | 16.70–22.74 |
| 1999 | 21.80 | 20.32–23.27 | 24.88 | 23.20–26.66 |
| 2000 | 22.73 | 21.22–24.23 | 25.30 | 23.64–27.06 |
| 2001 | 25.21 | 23.63–26.80 | 27.91 | 26.18–29.74 |
| 2002 | 25.76 | 24.17–27.36 | 27.99 | 26.27–29.80 |
| 2003 | 26.35 | 25.43–27.27 | 28.99 | 27.97–30.04 |
| 2004 | 28.11 | 27.16–29.06 | 30.20 | 29.17–31.26 |
| 2005 | 31.28 | 30.27–32.28 | 32.60 | 31.55–33.68 |
| 2006 | 32.33 | 31.30–33.35 | 33.02 | 31.98–34.10 |
| 2007 | 29.85 | 29.23–30.47 | 30.70 | 30.05–31.35 |
| 2008 | 35.43 | 34.76–36.11 | 35.54 | 34.86–36.24 |
| 2009 | 41.78 | 41.04–42.51 | 40.65 | 39.93–41.39 |
| 2010 | 44.84 | 44.08–45.60 | 42.79 | 42.06–43.53 |
| 2011 | 50.60 | 49.78–51.42 | 47.44 | 46.67–48.23 |
| 2012 | 54.97 | 54.12–55.82 | 50.56 | 49.77–51.35 |
| 2013 | 57.95 | 57.08–58.83 | 52.62 | 51.82–53.42 |
| 2014 | 61.76 | 60.86–62.66 | 55.08 | 54.27–55.89 |
| 2015 | 65.25 | 64.33–66.17 | 58.12 | 57.29–58.95 |
| 2016 | 61.52 | 60.62–62.41 | 54.01 | 53.22–54.81 |
| 2017 | 64.92 | 64.00–65.83 | 56.38 | 55.58–57.19 |
| 2018 | 62.55 | 61.65–63.45 | 53.64 | 52.86–54.43 |
| 2019 | 65.67 | 64.21–67.12 | 53.61 | 52.41–54.84 |

*Legend:* CIR = Crude Incidence Rate; ASIR = Age-Standardized Incidence Rate. Rates are expressed per 100,000 population. 95% CI = 95% Confidence Interval.

**Figure 2.**
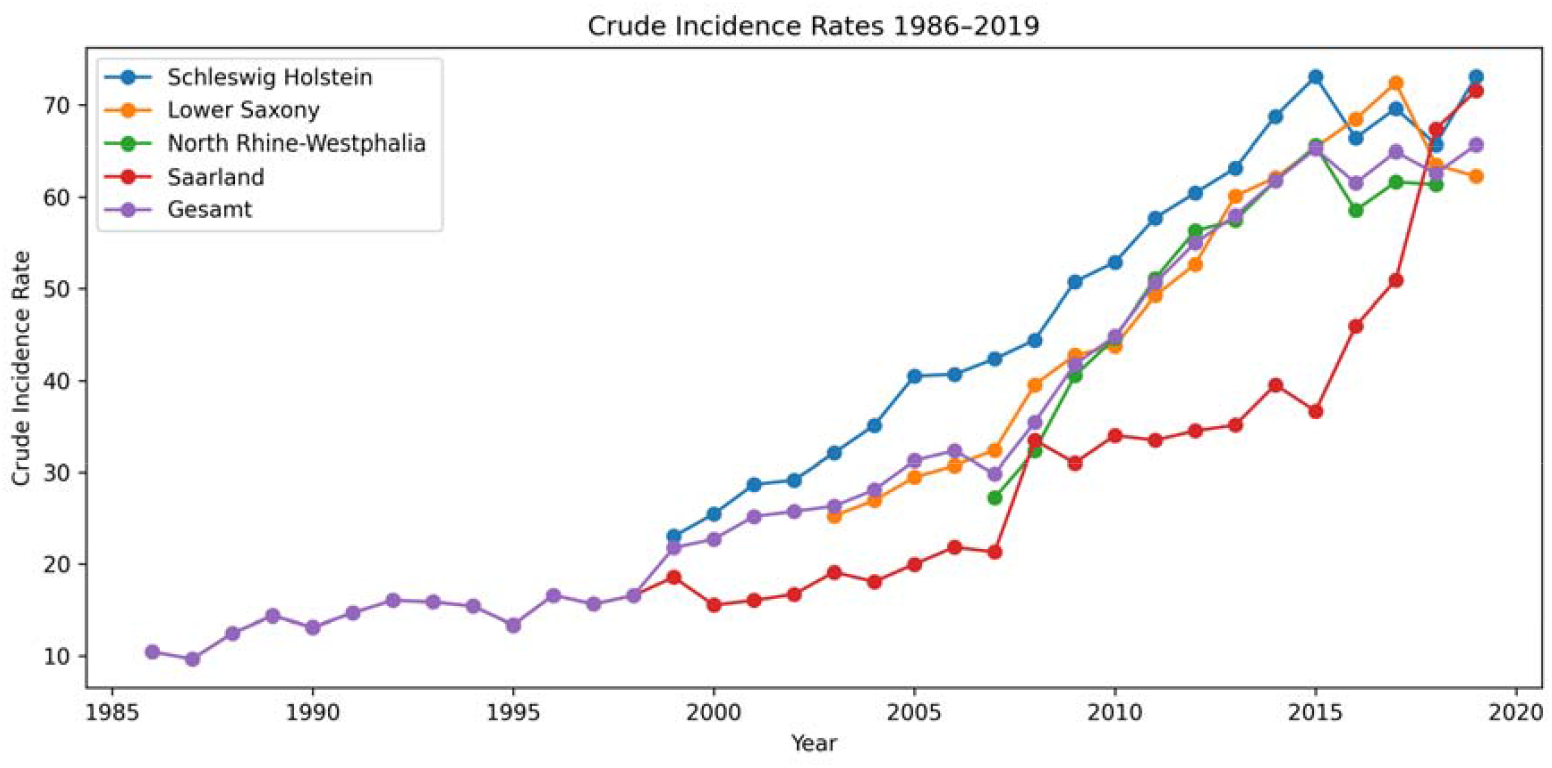
Crude Incidence Rates (CIR) in four federal states and total from 1986-2019.

**Figure 3.**
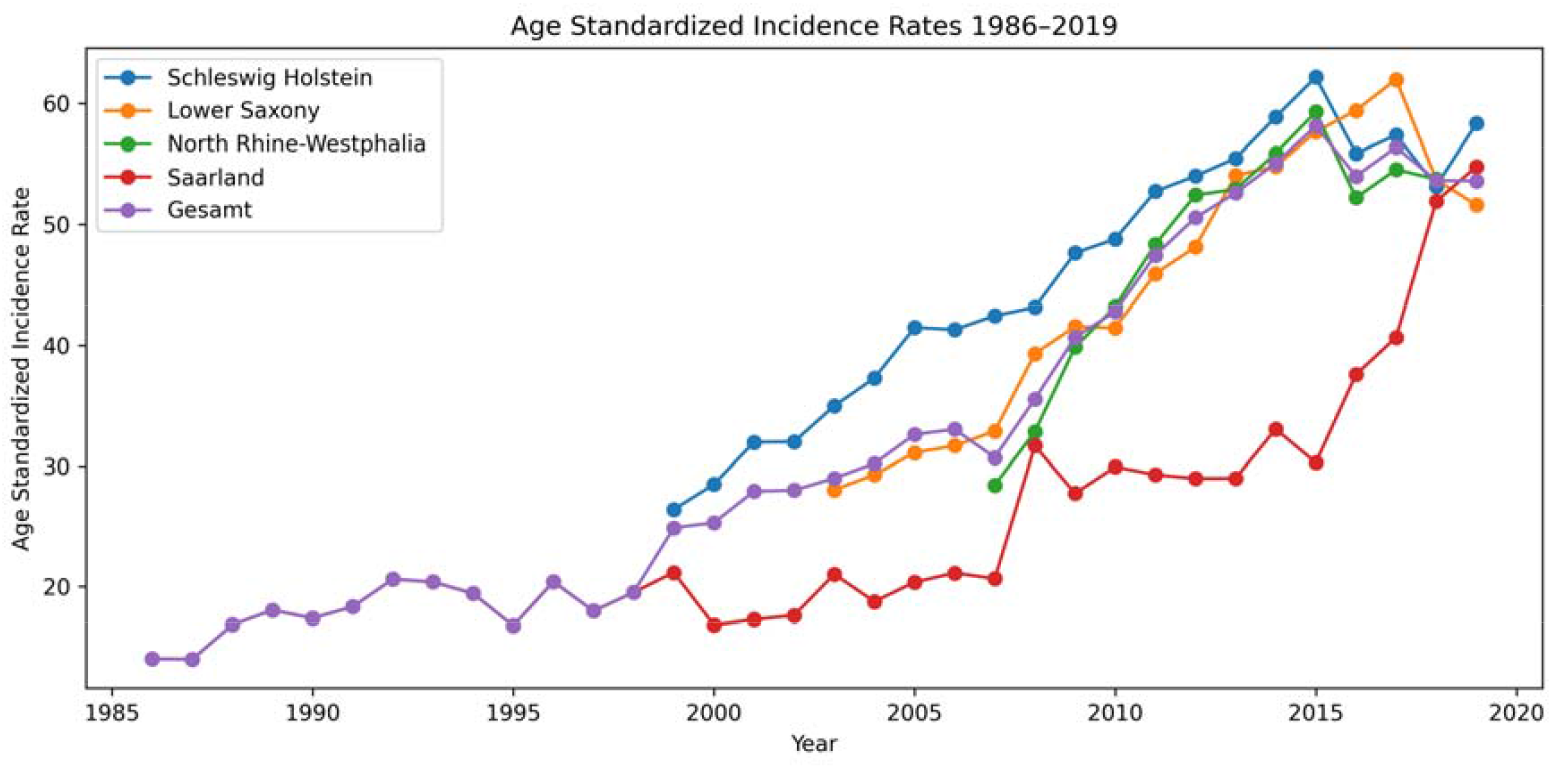
Age Standardized Incidence Rates (ASIR) in four federal states and total from 1986-2019.

The face (ICD-10 C44.3) was the most frequent tumor site, accounting for 40.4% of all cases, followed by the scalp/neck (C44.4) and upper extremities (C44.6). Men more often had tumors on the scalp, ear, and neck, while women more frequently developed tumors on the lower extremities. Tumors were more common on the right side of the body in both sexes (56.8%; p *<* 0.0001). The topographic distribution across ICD-10 categories differed significantly (*x_2_*= 868.59 for C44.0–C44.7; *x_2_*= 1005.80 for C44.0– C44.8; both p *< 0.000001*), with consistent patterns across federal states except for lower CIR values in Saarland. Full frequency, proportion, and incidence data are shown in Figure 4. T-stage data were available for 66.5% of cases (Table 3). Among these, 88.6% were T1, 9.1% T2, 1.8% T3, and 0.5% T4. Women had slightly more T1 tumors. T-stage distribution varied significantly by anatomical site (p *< 0.0001).*

**Table 3.**
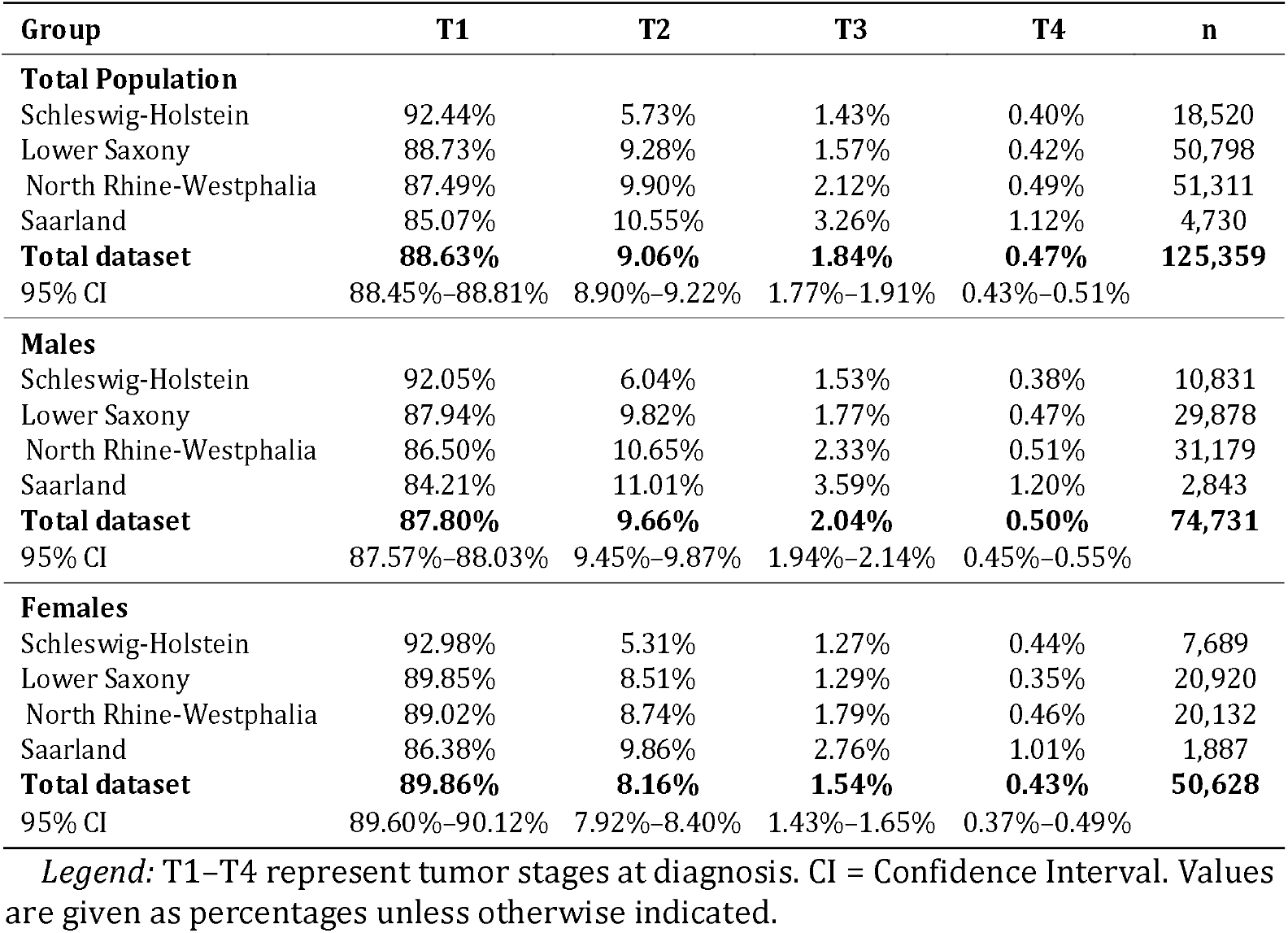
Distribution of Tumor Stages (T1–T4) by Federal State and Sex, Including 95% Confidence Intervals.

| Group | T1 | T2 | T3 | T4 | n |
| --- | --- | --- | --- | --- | --- |
| **Total Population** |  |  |  |  |  |
| Schleswig-Holstein | 92.44% | 5.73% | 1.43% | 0.40% | 18,520 |
| Lower Saxony | 88.73% | 9.28% | 1.57% | 0.42% | 50,798 |
| North Rhine-Westphalia | 87.49% | 9.90% | 2.12% | 0.49% | 51,311 |
| Saarland | 85.07% | 10.55% | 3.26% | 1.12% | 4,730 |
| **Total dataset** | **88.63%** | **9.06%** | **1.84%** | **0.47%** | **125,359** |
| 95% CI | 88.45%–88.81% | 8.90%–9.22% | 1.77%–1.91% | 0.43%–0.51% |  |
| **Males** |  |  |  |  |  |
| Schleswig-Holstein | 92.05% | 6.04% | 1.53% | 0.38% | 10,831 |
| Lower Saxony | 87.94% | 9.82% | 1.77% | 0.47% | 29,878 |
| North Rhine-Westphalia | 86.50% | 10.65% | 2.33% | 0.51% | 31,179 |
| Saarland | 84.21% | 11.01% | 3.59% | 1.20% | 2,843 |
| **Total dataset** | **87.80%** | **9.66%** | **2.04%** | **0.50%** | **74,731** |
| 95% CI | 87.57%–88.03% | 9.45%–9.87% | 1.94%–2.14% | 0.45%–0.55% |  |
| **Females** |  |  |  |  |  |
| Schleswig-Holstein | 92.98% | 5.31% | 1.27% | 0.44% | 7,689 |
| Lower Saxony | 89.85% | 8.51% | 1.29% | 0.35% | 20,920 |
| North Rhine-Westphalia | 89.02% | 8.74% | 1.79% | 0.46% | 20,132 |
| Saarland | 86.38% | 9.86% | 2.76% | 1.01% | 1,887 |
| **Total dataset** | **89.86%** | **8.16%** | **1.54%** | **0.43%** | **50,628** |
| 95% CI | 89.60%–90.12% | 7.92%–8.40% | 1.43%–1.65% | 0.37%–0.49% |  |

*Legend:* T1–T4 represent tumor stages at diagnosis. CI = Confidence Interval. Values are given as percentages unless otherwise indicated.

**Figure 4.**
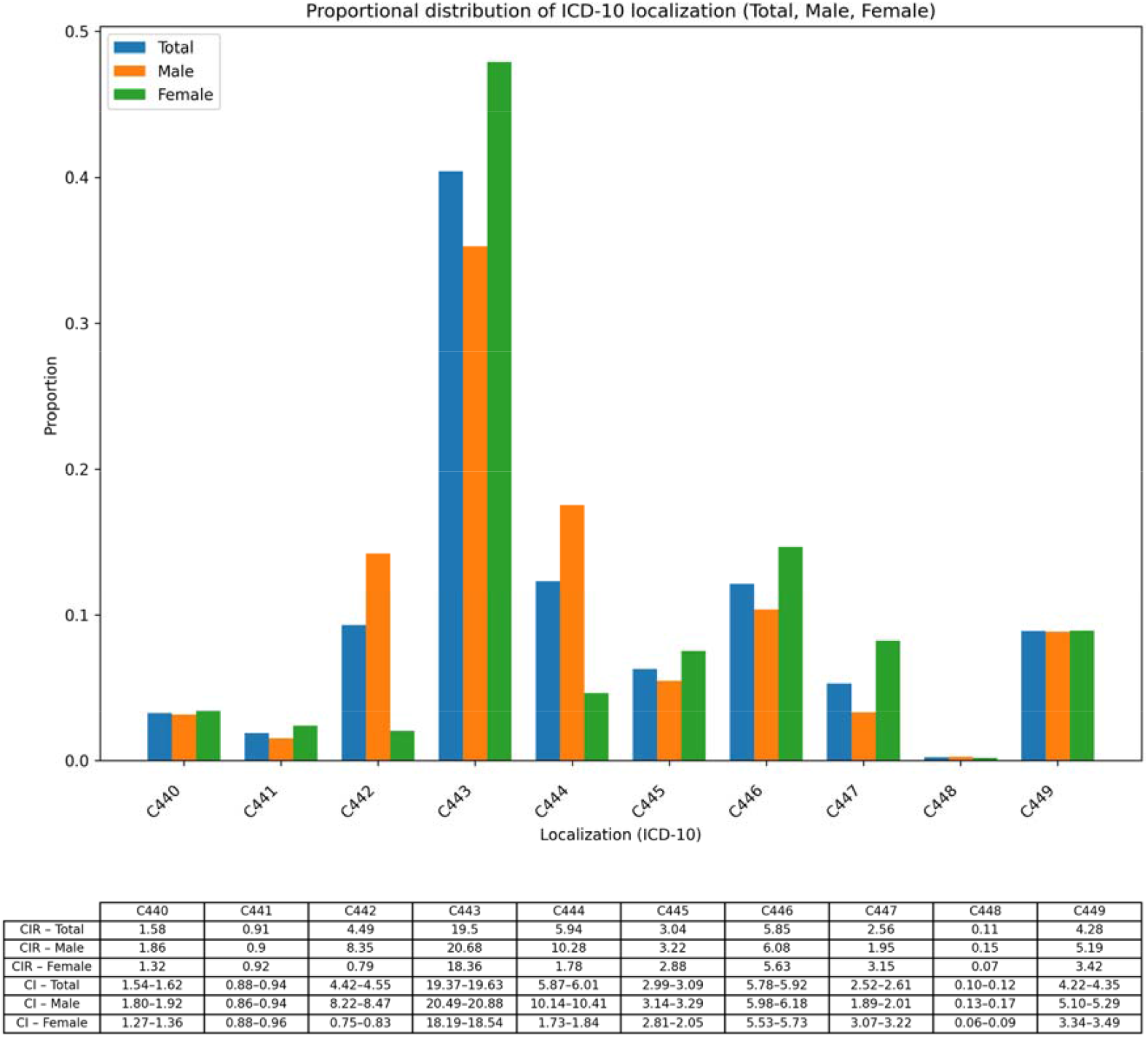
ICD-10 distribution of primary cSCC in Germany for males, females and total. Proportional Frequencies are plotted for each ICD-10 code. Crude Incidence Rates (CIR) and Confidence Intervals (CI) are provided in the data frame below.

Histological subtype data were available for 207,515 cases. Most were cSCC NOS (56.8%) or keratinizing cSCC (34.6%). Subtype distribution varied by state. More aggressive subtypes such as large-cell non-keratinizing- and spindle cell type cSCC were associated with higher T-stages.

## Discussion

This study provides a comprehensive epidemiological assessment of the first invasive cSCC in Germany, drawing on more than three decades of high-quality cancer registry data. With ∼ 214.000 patients the datasets magnitude exceeds the previously largest analysis on ∼145.000 cases in Europe ^7^. The sustained rise in incidence, particularly among women, aligns with international trends and may reflect cumulative UV exposure, ageing populations, and increased diagnostic scrutiny ^2,7,8,10^. The sharper increase in female incidence, despite a higher absolute burden in men, may be due to longevity, behavioral factors, or improved access to dermatological care.

From 2015 onwards, age-standardized incidence rates plateaued or declined in three of the four participating federal states. While this may suggest a stabilization in real incidence, alternative explanations such as saturation of detection, regional changes in reporting, or data processing delays cannot be excluded. Saarland’s delayed increase to match national levels highlights the heterogeneity of registry dynamics and underlines the importance of regional data quality.

Sex-specific patterns of anatomical site were consistent with known behavioral and anatomical differences ^11^. Men showed a higher prevalence of tumors on the scalp, ear, and neck, likely due to shorter hair and increased cumulative UV exposure. Women more frequently developed cSCC on the lower extremities, a pattern reported previously and possibly influenced by grooming practices and clothing. Interestingly, facial tumors were equally frequent in both sexes, suggesting general UV vulnerability of this highly exposed region.

Despite this similarity, there was a distinctive pattern when males or females develop cSCC. Men develop cSCC significantly earlier than women. From 60 years of age onwards, men’s risk to develop cSCC is at least twice as much compared to females. The predominance of T1 tumors in our cohort supports early detection, particularly in visible and sun-exposed anatomical regions such as the face and scalp. This distribution corresponds with clinical experience, where cSCC is often diagnosed at an early stage during routine dermatological examinations. However, interpretation of T-stage distributions in cancer registry data requires caution. T-stage was missing in 34.5% of cases, limiting the representativeness of staging data. Accurate classification between T1 and T2 depends primarily on clinical parameters, especially lesion diameter, which is often incompletely documented. Histological parameters such as tumor thickness *> 6 mm, pe*rineural invasion, infiltration beyond subcutaneous fat, or bone involvement typically indicate T3 or higher. Therefore, the high proportion of T1 tumors in our cohort likely reflects both true early-stage detection and limitations in the availability of clinical staging data.

The regional and sex-specific T-stage distributions observed in our cohort further support the heterogeneity of cSCC presentation and documentation. While Schleswig-Holstein showed the highest overall T1 proportion (92.4%), Saarland consistently reported lower T1 rates and a markedly higher proportion of advanced tumors, particularly among men. Male patients from Saarland had a combined T3–T4 rate of 4.8%, compared to 3.8% in North Rhine-Westphalia and less than 2% in Schleswig-Holstein. These differences may reflect delayed clinical detection, disparities in access to specialist care, or inconsistencies in the integration of clinical and pathological staging data. Female patients across all regions displayed a more favorable T-stage profile, with higher proportions of T1 and fewer advanced tumors. Whether these findings reflect biological differences, health-seeking behaviors, or diagnostic practices requires further investigation.

Histological subtypes showed expected dominance of keratinizing and NOS variants. The marked variation in subtype coding between federal states, especially the inverse correlation between NOS frequency and the reporting of less common subtypes, suggests differences in diagnostic precision and may reflect disparities in pathology practice or registry completeness. Subtypes such as large-cell non-keratinizing and spindle cell carcinoma were associated with higher T-stages, underlining the importance of histological classification for risk stratification and treatment planning. The current registry framework captures only the first primary tumor per patient, omitting subsequent or recurrent cSCCs. This limitation considerably underestimates clinical workload, particularly in patients with actinic field cancerization, where multiple primaries are common. Data from the Netherlands suggest that more than 30% of cSCC cases are second or later tumors ^12^. Moreover, the exclusion of carcinoma in situ such as Bowen’s disease or the precursor lesion actinic keratosis from registry data omits a significant component of the therapeutic burden, as field-directed treatment of such precursor lesions is central to cSCC prevention ^6,13,14^.

In summary, this nationwide registry-based analysis confirms that cSCC is a highly prevalent and clinically relevant malignancy, with rising incidence in Germany over recent decades. The observed trends reveal distinct sex- and site-specific patterns and considerable variation in tumor staging and histological classification. The predominance of T1 tumors suggests effective early detection, particularly for visible anatomical regions, but is offset by incomplete staging data and the exclusion of multiple and precursor lesions from routine surveillance. Strengthening registry protocols to include multiple primaries and early-stage conditions, and ensuring better integration of clinical staging information, will be essential to improve epidemiological accuracy and support targeted prevention strategies in dermatologic oncology.

## Data Availability

Data used in this research is available upon a utilization request at the Robert Koch Institute.

